# Prognostic Predictors After Bone Marrow-Derived Mononuclear Cell Implantation in No-Option Chronic Limb-Threatening Ischemia Patients with Atherosclerotic Lower Extremity Artery Disease

**DOI:** 10.1101/2023.10.25.23297576

**Authors:** Hirofumi Kawamata, Kenji Yanishi, Jun Yoshimura, Takaaki Ozawa, Daiki Goto, Yusuke Hori, Ayumu Fujioka, Keisuke Shoji, Arito Yukawa, Satoaki Matoba

## Abstract

**Background:** Previous studies have reported the efficacy and safety of therapeutic angiogenesis through bone marrow-derived mononuclear cell (BM-MNC) implantation in patients with no-option critical limb-threatening ischemia (CLTI) from atherosclerotic lower extremity artery disease (LEAD). However, uncertain clinical prognostic factors impact treatment outcomes.

**Methods:** In this retrospective, single-center, observational study, we assessed the long-term prognosis post-treatment. Primary endpoints included the long-term prognosis of BM-MNC implantation and factors influencing 1-year outcomes.

**Results:** A total of 92 limbs in 84 patients were analyzed in the final cohort. The mean age was 67 years, and 65% were male. The 5- and 10-year overall survival rates were 50.0% and 31.0%, respectively, while the 5- and 10-year amputation-free survival rates were 37.6% and 23.3%, respectively. Multivariate logistic analysis linked all-cause mortality to an age ≥70 years, hemodialysis, smoking, and a controlling nutrition status score ≥5. Major amputation or mortality was associated with male gender, hemodialysis, and C-reactive protein levels ≥3.0 mg/dL. No adverse events were associated with therapeutic angiogenesis.

**Conclusions:** These findings endorse the feasibility and safety of BM-MNC implantation for patients with no-option CLTI due to atherosclerotic LEAD. Moreover, the study highlights the significance of several prognostic factors, including advanced age, hemodialysis, smoking, and inflammatory markers, in influencing the long-term outcomes of this treatment.

**Clinical Perspective:** *What is new?:* This study shows a new scoring model of therapeutic angiogenesis using autologous bone marrow-derived mononuclear cell implantation in patients or their limbs with no-option chronic limb-threatening ischemia (CLTI) attributed to atherosclerotic lower extremity artery disease (LEAD). High age, hemodialysis, smoking, malnutrition, ambulatory, and inflammatory response are identified as prognostic factors. A scoring formula, developed through these factors, effectively identifies a group with a favorable long-term prognosis in both patients and limbs. The counts of bone marrow-derived mononuclear cells and CD34 surface antigen-positive cells are found to have a significant relationship with a 1-year prognosis in both patients and limbs.

*What are the clinical implications?:* This study demonstrates the feasibility and safety of bone marrow-derived mononuclear cell implantation among patients with no-option CLTI patients resulting from LEAD. This scoring model will help us predict the long-term prognosis of patients and their affected limbs treated by bone marrow-derived mononuclear cell implantation. These results also provide valuable information for choosing a personalized treatment plan for each patient.

## INTRODUCTION

Chronic limb-threatening ischemia (CLTI) is the most severe clinical manifestation of lower extremity artery disease (LEAD) [1, 2]. In patients with CLTI, the risk of limb amputation and mortality is high. This risk persists despite optimal medical treatment, wound care, and revascularization approaches, including endovascular therapy (EVT) and bypass surgery [3–9]. Despite receiving conventional standard treatments, some patients with CLTI do not exhibit improvements regarding necrosis and infectious diseases. In such cases, the option of limb amputation must be considered to save their lives. Therapeutic angiogenesis using the implantation of autologous bone marrow-derived mononuclear cells (BM-MNCs) is indicated for those patients with CLTI who poorly respond to conventional treatments (no-option CLTI). This therapeutic approach is conducted in accordance with the Act on Securing Safety of Regenerative Medicine. This treatment entails administering BM-MNCs from the patient’s own bone marrow fluid to the ischemic limb, aiming to stimulate the CD34 surface antigen-positive (CD34+) cells contained in the BM-MNCs to facilitate the formation of new blood vessels and improve the microcirculation in ischemic limbs [10–13]. This treatment primarily targets CLTI resulting from atherosclerotic LEAD, thromboangiitis obliterans (TAO), and collagen disease-associated vasculitis (CDV). Numerous clinical studies have reported the effectiveness and safety of BM-MNC implantation for CLTI [13–23]. However, limb salvage and survival rates after BM-MNC implantation in CLTI caused by atherosclerotic LEAD are worse than those in CLTI caused by the other two diseases (non-atherosclerotic LEAD) [13, 17, 18, 20]. In atherosclerotic LEAD, this is mainly attributed to the higher prevalence of background factors, such as hemodialysis (HD), diabetes mellitus (DM), and cardiovascular disease. These factors have a more adverse impact on the prognosis of the patients and their limbs compared to non-atherosclerotic LEAD. However, in previous studies, the background factors that influence the long-term prognosis of BM-MNC implantation were not fully examined. This study aimed to elucidate the long-term outcomes of patients with atherosclerotic LEAD-derived no-option CLTI after BM-MNC implantation and to identify prognostic factors that influence these outcomes. The study also sought to build a model to predict the prognosis of patients and affected limbs treated by BM-MNC implantation.

## METHODS

### Study design

This was a single-center, retrospective, observational study. A survey was conducted to assess the long-term prognosis of patients with atherosclerotic LEAD-derived no-option CLTI (both patients and affected limbs) who underwent therapeutic angiogenesis using BM-MNC implantation at our hospital between January 2010 and December 2022. Information on target cases continuing treatment at our hospital was collected from the medical records. For cases that were transferred to other facilities, where the outcome could not be determined at our hospital, outcome questionnaires were sent to the respective facilities, and responses were obtained and analyzed.

### Participants

Participants who underwent BM-MNC implantation were individuals diagnosed with no-option CLTI classified as Fontaine III or IV, of both sexes (male or female), and aged between 20 and 80 years. Patients with no-option CLTI were defined as individuals who were refractory to or not indicated for any other conventional treatment options (e.g., medical therapy, wound care, sympathetic ganglion block, and revascularization by EVT or surgery) that could potentially lead to amputation. Before BM-MNC implantation, we assessed participants to determine if they met any of the exclusion criteria. The exclusion criteria included untreated coronary artery disease (CAD) or cerebrovascular disease (CVD), clinical or laboratory signs of chronic or acute inflammation, a current malignant tumor condition, DM with untreated retinopathy, the possibility of pregnancy, or a lack of informed consent.

### Procedure

BM-MNC implantation was performed under general anesthesia (Figure 1). First, approximately 600–800 mL of bone marrow fluid was collected from both iliac bones. Subsequently, the collected bone marrow fluid was separated using a blood component separator and concentrated to approximately 60–80 mL, containing >0.5×10^9^ BM-MNCs. The BM-MNCs were evenly injected into the skeletal muscles of the target leg(s) below the knee. After BM-MNC implantation, patients were discharged from our hospital after 7–10 days, and they continued to receive regular follow-up care while undergoing conventional treatments for CLTI. The numbers of BM-MNCs and CD34+ cells were measured using a portion of the collected bone marrow fluid.

**Figure 1.**
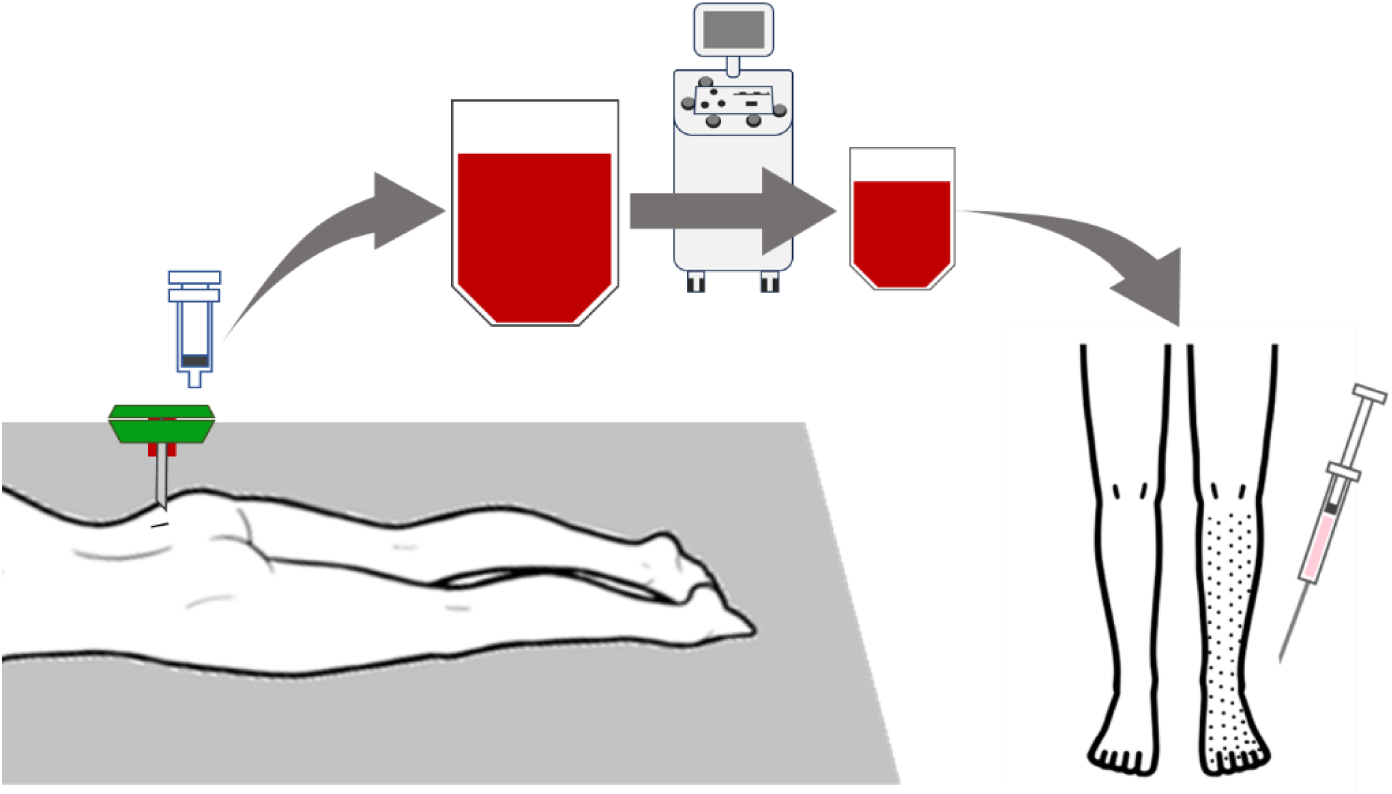
Schematic representation of the BM-MNC implantation procedure. BM-MNC implantation is performed under general anesthesia. Bone marrow fluid is collected from both iliac bones. Then, the collected bone marrow fluid is separated using a blood component separator and concentrated. The concentrated fluid contains BM-MNC, which are injected into the skeletal muscles of the target leg(s) below the knee. BM-MNC, bone marrow-derived mononuclear cell

### Outcomes

The primary endpoints of this study included the long-term prognosis of BM-MNC implantation and the factors related to the 1-year prognosis of this treatment. A prediction scoring model based on these factors was also developed. Long-term prognosis was evaluated using the overall survival (OS) and major amputation-free survival (AFS) rates. Factors associated with all-cause death (AD) and major amputation or death (MA/D) within 1 year of BM-MNC implantation were analyzed. The secondary endpoints included adverse events within 6 months of BM-MNC implantation, factors related to major/minor amputation within 6 months of BM-MNC implantation, and the relationship between AD and MA/D within 1 year after BM-MNC implantation, together with the number of BM-MNCs and CD34+ cells. The following adverse events were included: AD, CAD, CVD, heart failure, aortic disease, MA/D, major amputation, minor amputation, and revascularization.

### Ethics statement

BM-MNC implantation was approved by the Ministry of Health, Labour and Welfare after being approved by the Certified Committee for Regenerative Medicine in the Kyoto Prefectural University of Medicine in 2015 (approval number, NA8150008). In addition, this long-term outcome survey was approved by the Ethics Committee of Kyoto Prefectural University of Medicine (approval number, ERB-C-2264). The study was also registered with the University Hospital Medical Information Network (UMIN) Clinical Trials Registry (ID: UMIN000046192). In accordance with the ethical guidelines of the Declaration of Helsinki, written informed consent was obtained from all patients.

### Statistical analysis

Data were aggregated for each patient and affected limb, and these were defined as patient and limb data, respectively. Patient data were utilized to analyze OS and AD, and limb data were utilized to analyze AFS and MA/D. Continuous variables are expressed as mean ± standard deviation. Logistic regression analysis was used to analyze variables related to prognosis, and the results are expressed as odds ratios (ORs) and 95% confidence intervals (95% CIs). Multivariate logistic regression analysis used a stepwise method, and the threshold for inclusion and exclusion in the model was set at *P* = 0.25. The multivariate logistic analysis results were used to derive a formula for predicting the risk of AD and MA/D 1 year after BM-MNC implantation. The Kaplan–Meier method was used to analyze time-dependent outcomes, including OS and AFS. All *P*-values were calculated using the Wald test, and *P* < 0.05 was considered statistically significant. JMP 14^®^ (SAS Institute Inc., Cary, NC, USA) was used for statistical analysis.

## RESULTS

### Study flowchart

This study included 139 patients who underwent BM-MNC implantation at our hospital. A total of 55 patients were excluded for the following reasons: 5 patients did not undergo lower extremity treatment, 14 had received the same treatment in the past, 31 also had CLTI with non-atherosclerotic pathology, and 5 were unavailable for follow-up observations. A total of 92 limbs in 84 patients were ultimately included in the analysis of this study (Figure 2).

**Figure 2.**
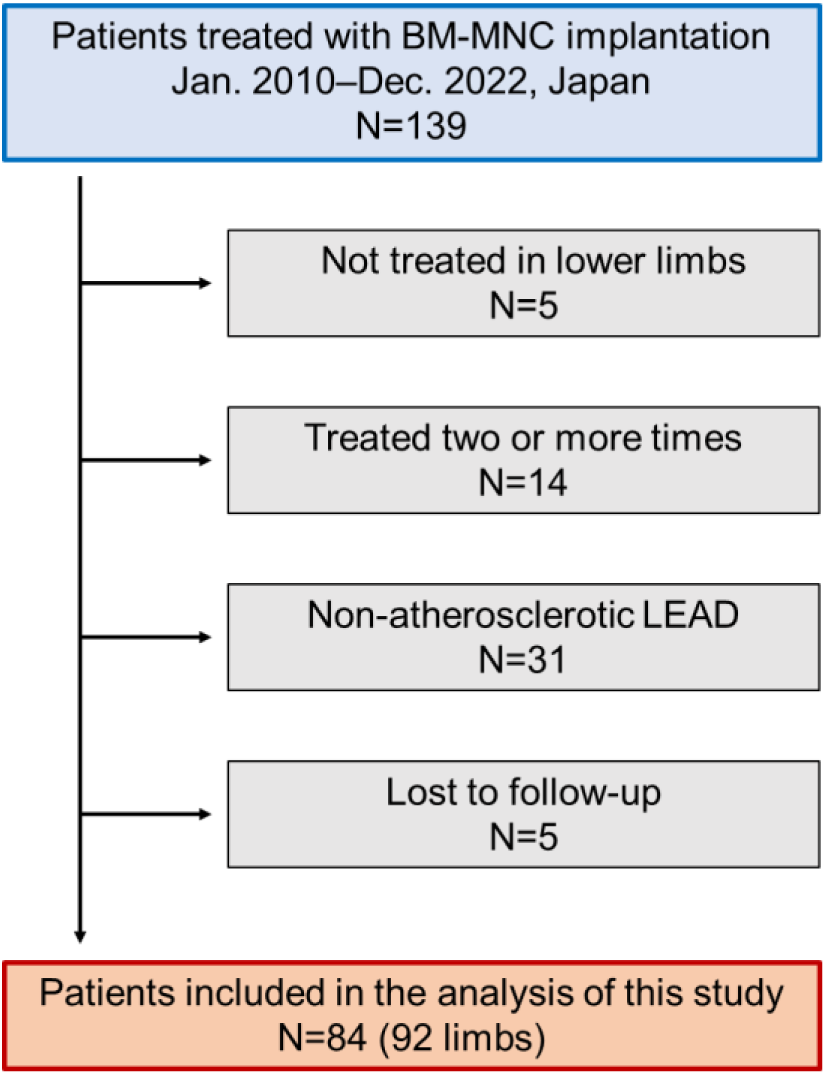
Flowchart illustrating the patient selection process. BM-MNC, bone marrow-derived mononuclear cell; LEAD, lower extremity artery disease.

### Baseline characteristics

The baseline characteristics are shown in Table 1. The average age of the patients was 67 years. All patients were Japanese, and 55 were males (65%). Of the patients, 48 (57%), 36 (43%), and 21 (25%) had DM, HD, and heart failure, respectively. Among the patients, 54 (64%) were ambulatory, 26 (31%) had a moderately impaired nutritional status (defined in this study as a Controlling Nutritional Status [CONUT] score of ≥ 5), and 56 (67%) had a history of smoking. The BM-MNC implantation procedure was successful in all patients, and no intraoperative adverse events were observed. The average amount of bone marrow fluid collected from patients per body weight was 13 mL/kg. The average counts of BM-MNCs and CD34+ cells administered per patient were 2.1×10^9^ and 3.5×10^7^, respectively. Among the affected limbs, the treatment was administered to the right lower extremity in 51 (55%) limbs, with 68 (74%) limbs categorized as Fontaine stage IV, and 20 (22%) limbs classified as Rutherford category 6.

**Table 1.**
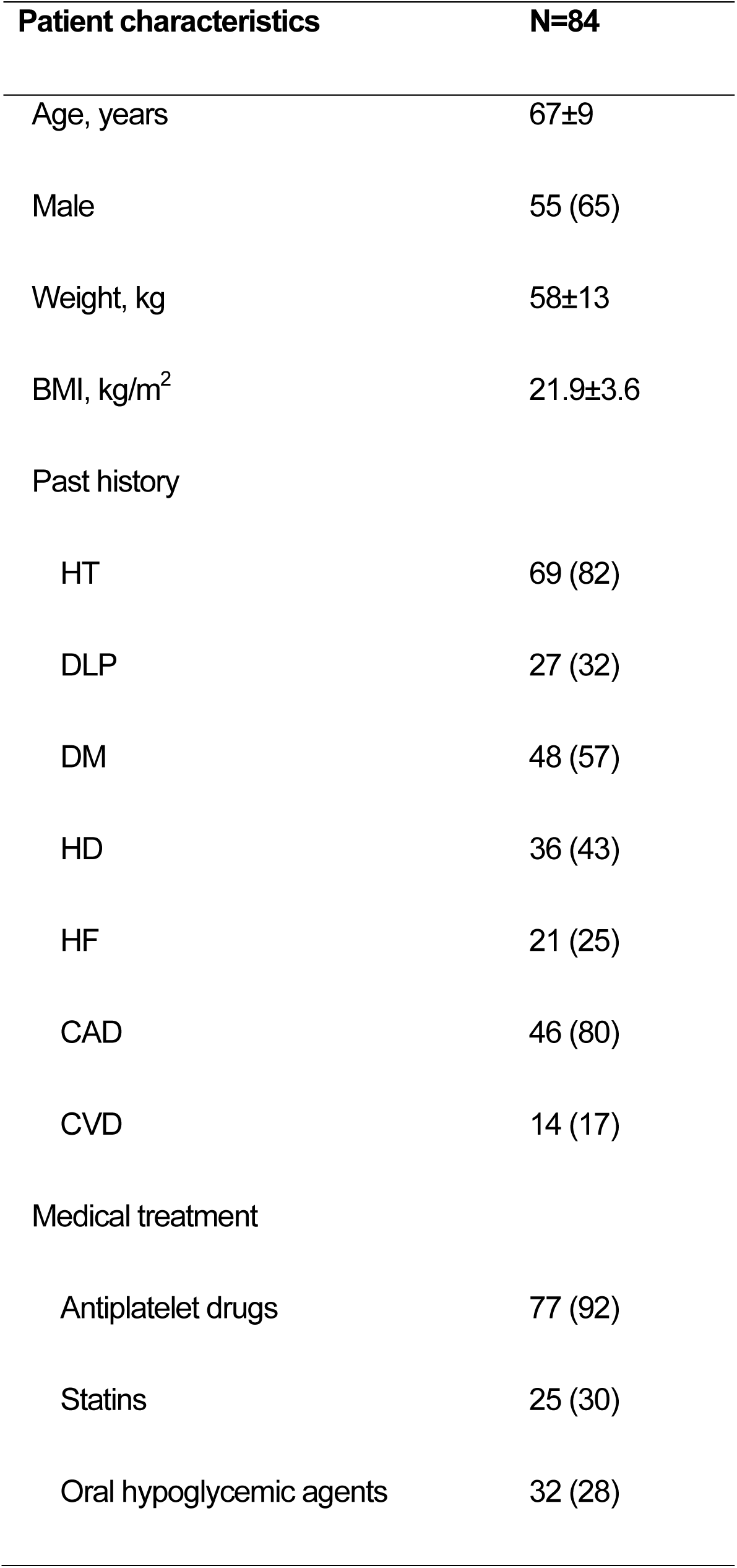
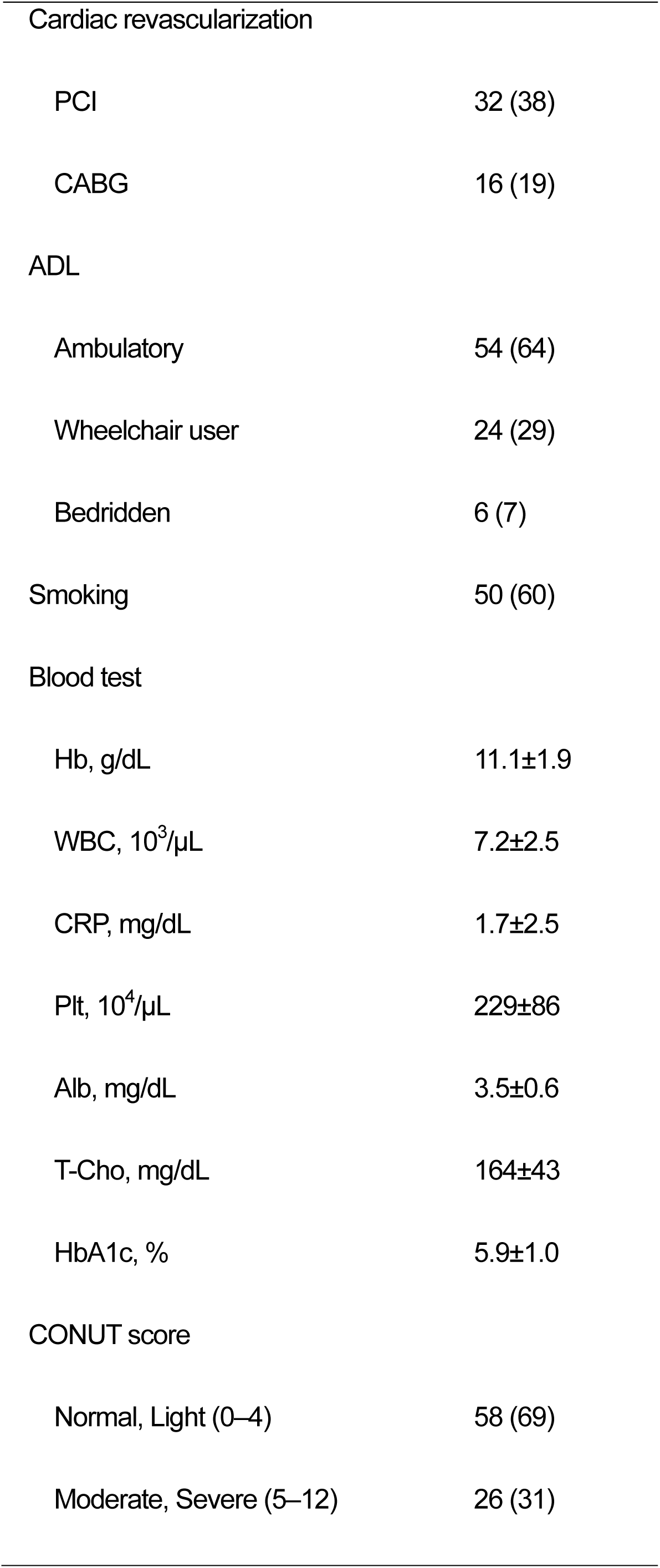
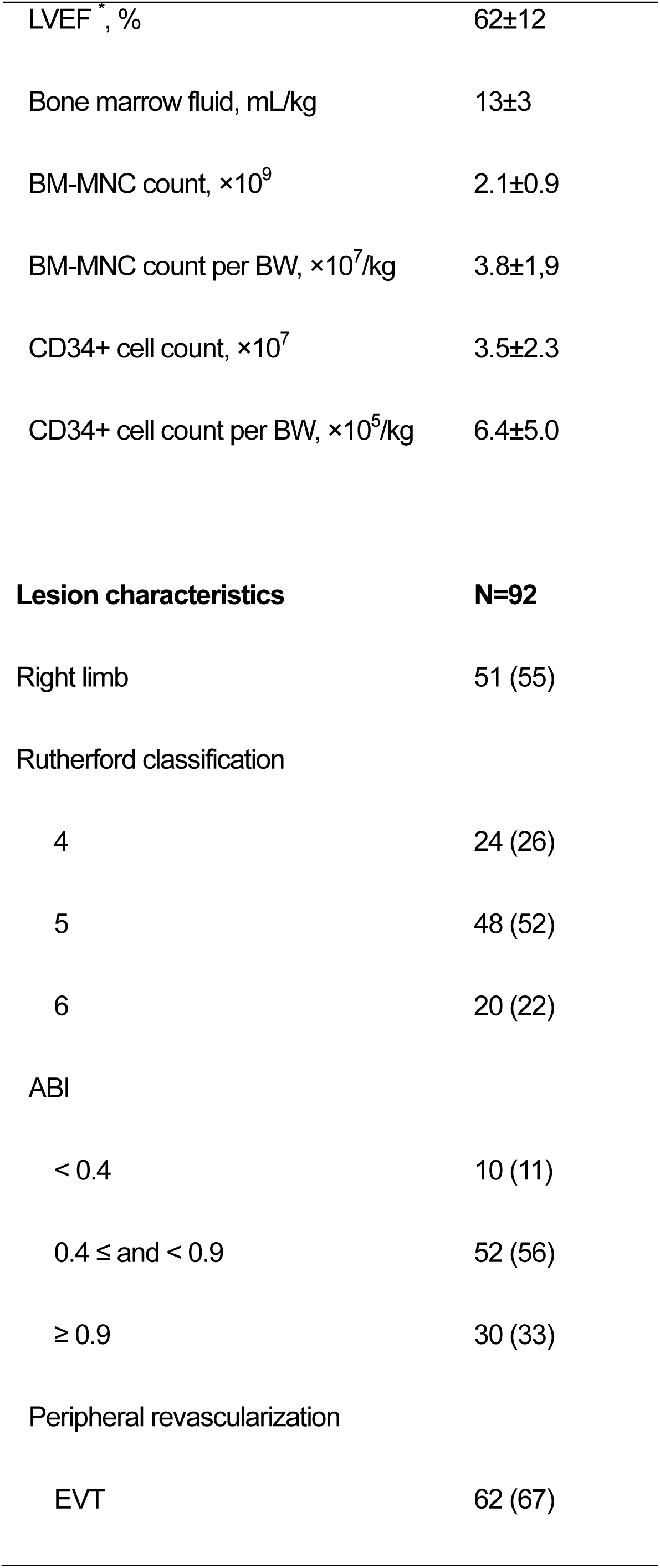
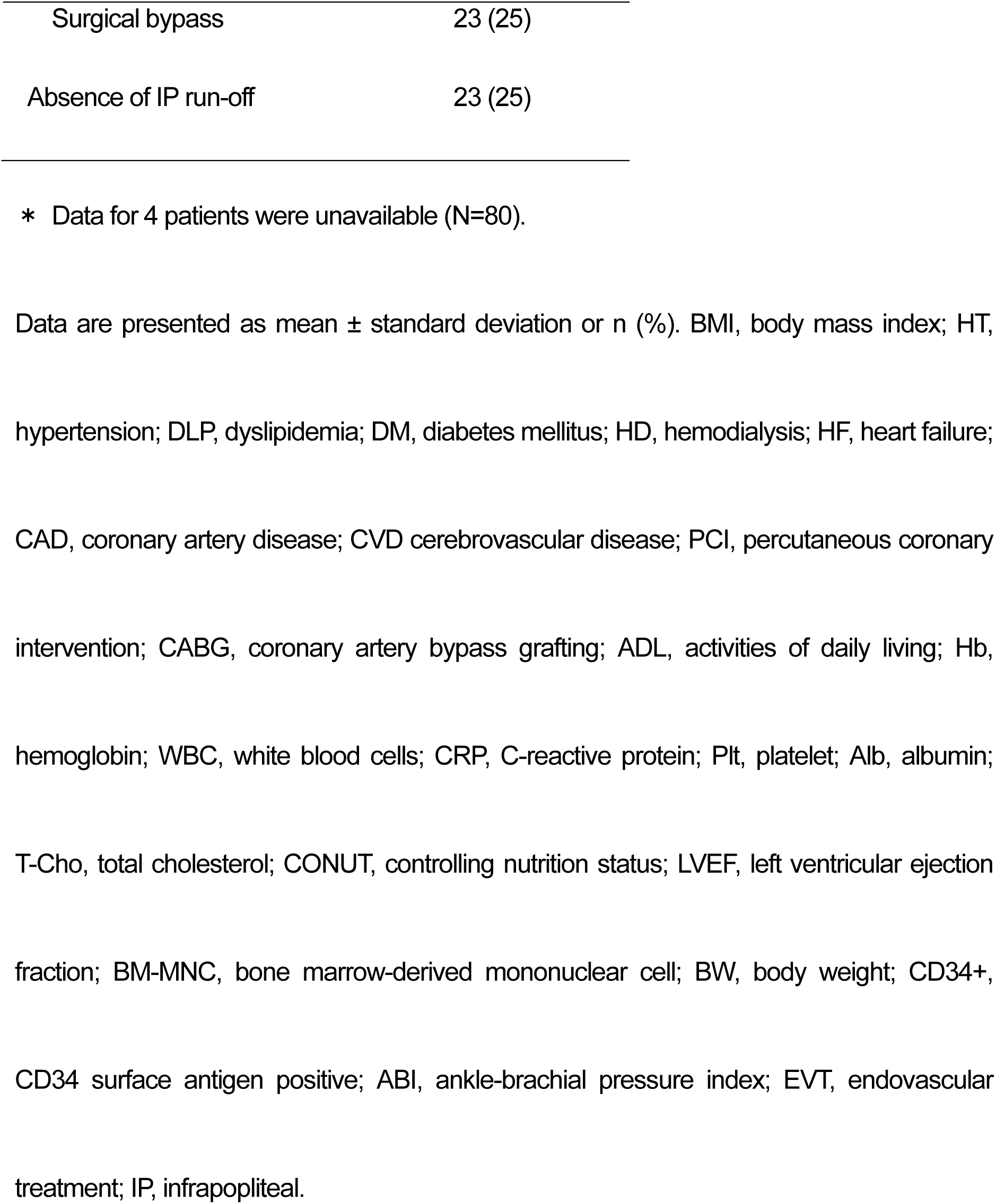
Baseline characteristics of study participants.

| **Patient characteristics** | **N=84** |
| --- | --- |
| Age, years | 67±9 |
| Male | 55 (65) |
| Weight, kg | 58±13 |
| BMI, kg/m<sup>2</sup> | 21.9±3.6 |
| Past history |  |
| HT | 69 (82) |
| DLP | 27 (32) |
| DM | 48 (57) |
| HD | 36 (43) |
| HF | 21 (25) |
| CAD | 46 (80) |
| CVD | 14 (17) |
| Medical treatment |  |
| Antiplatelet drugs | 77 (92) |
| Statins | 25 (30) |
| Oral hypoglycemic agents | 32 (28) |
| Cardiac revascularization |  |
| PCI | 32 (38) |
| CABG | 16 (19) |
| ADL |  |
| Ambulatory | 54 (64) |
| Wheelchair user | 24 (29) |
| Bedridden | 6 (7) |
| Smoking | 50 (60) |
| Blood test |  |
| Hb, g/dL | 11.1±1.9 |
| WBC, 10<sup>3</sup>/μL | 7.2±2.5 |
| CRP, mg/dL | 1.7±2.5 |
| Plt, 10<sup>4</sup>/μL | 229±86 |
| Alb, mg/dL | 3.5±0.6 |
| T-Chol, mg/dL | 164±43 |
| HbA1c, % | 5.9±1.0 |
| CONUT score |  |
| Normal, Light (0–4) | 58 (69) |
| Moderate, Severe (5–12) | 26 (31) |
| LVEF *, % | 62±12 |
| Bone marrow fluid, mL/kg | 13±3 |
| BM-MNC count, ×10<sup>9</sup> | 2.1±0.9 |
| BM-MNC count per BW, ×10<sup>7</sup>/kg | 3.8±1,9 |
| CD34+ cell count, ×10<sup>7</sup> | 3.5±2.3 |
| CD34+ cell count per BW, ×10<sup>5</sup>/kg | 6.4±5.0 |
| **Lesion characteristics** | **N=92** |
| Right limb | 51 (55) |
| Rutherford classification |  |
| 4 | 24 (26) |
| 5 | 48 (52) |
| 6 | 20 (22) |
| ABI |  |
| < 0.4 | 10 (11) |
| 0.4 ≤ and < 0.9 | 52 (56) |
| ≥ 0.9 | 30 (33) |
| Peripheral revascularization |  |
| EVT | 62 (67) |
| Surgical bypass | 23 (25) |
| Absence of IP run-off | 23 (25) |

\* Data for 4 patients were unavailable (N=80).
Data are presented as mean ± standard deviation or n (%). BMI, body mass index; HT, hypertension; DLP, dyslipidemia; DM, diabetes mellitus; HD, hemodialysis; HF, heart failure; CAD, coronary artery disease; CVD cerebrovascular disease; PCI, percutaneous coronary intervention; CABG, coronary artery bypass grafting; ADL, activities of daily living; Hb, hemoglobin; WBC, white blood cells; CRP, C-reactive protein; Plt, platelet; Alb, albumin; T-Cho, total cholesterol; CONUT, controlling nutrition status; LVEF, left ventricular ejection fraction; BM-MNC, bone marrow-derived mononuclear cell; BW, body weight; CD34+, CD34 surface antigen positive; ABI, ankle-brachial pressure index; EVT, endovascular treatment; IP, infrapopliteal.

### Long-term outcomes

Figure 3 displays the Kaplan–Meier curves illustrating the OS and AFS rates. Among the 84 patients, the OS rates at 6 months, 1 year, 5 years, and 10 years were 90.5%, 86.8%, 50.0%, and 31.0%, respectively (Figure 3A). The median follow-up period was 29.5 months. During the observation period, 34 (40%) patients died. After excluding 24 cases with unknown causes of death, 4 (17%) patients were found to have died from cardiovascular disease, with heart failure identified as the cause of death in all cases. Among the 92 affected limbs, the AFS rates at 6 months, 1 year, 5 years, and 10 years were 77.2%, 68.4%, 37.6%, and 23.3%, respectively (Figure 3B). The median follow-up period was 21 months.

**Figure 3.**
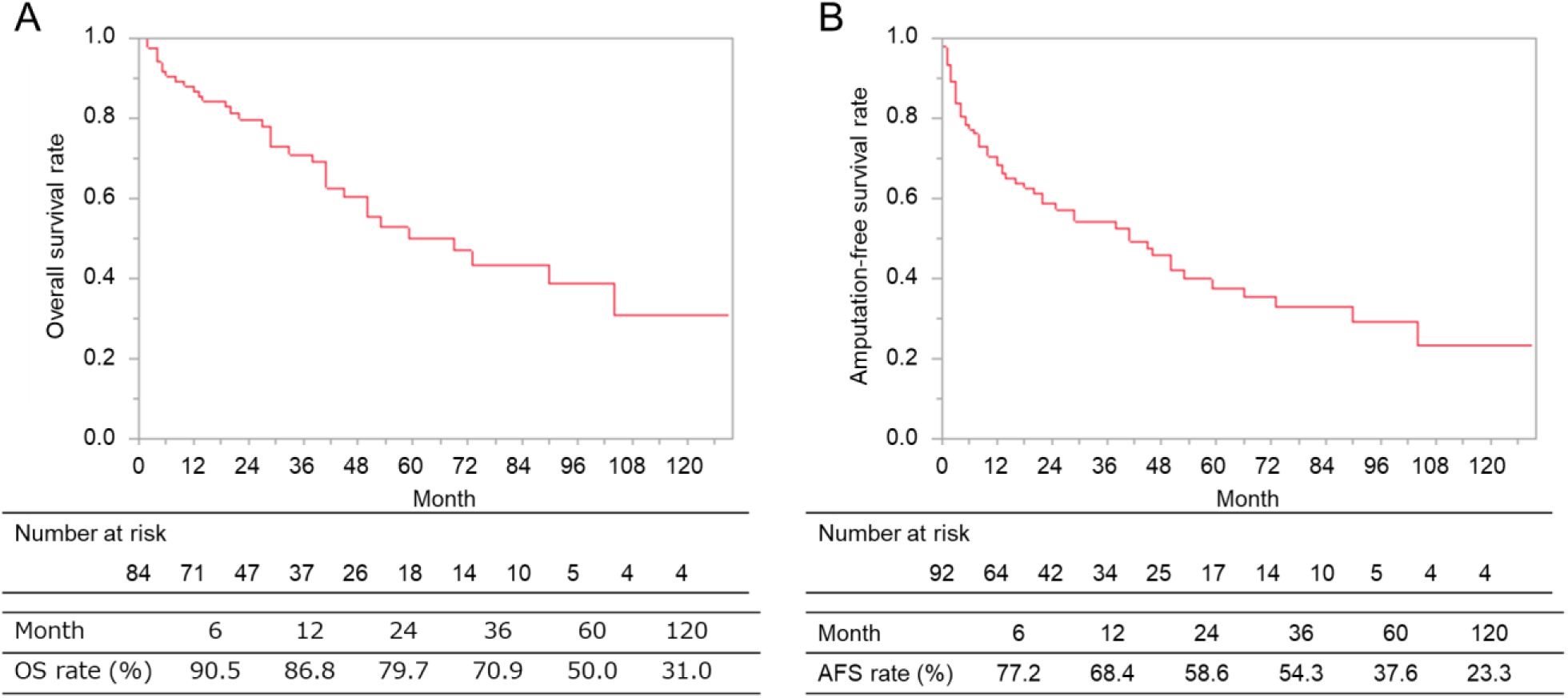
Long-term outcomes of OS (A) and AFS (B) rates. The OS rate was determined based on data from 84 patients, and the AFS rate was calculated using data from 92 limbs. OS, overall survival; AFS, amputation-free survival.

### Prognostic factors

Table 2A presents the results of a logistic regression analysis of variables related to AD within 1 year. In the univariate analysis, HD (OR: 7.62, 95% CI: 1.53–38.0, *P* < 0.05) was significantly associated with AD. Stepwise multivariate analysis showed that an age of ≥ 70 years (OR: 6.09, 95%CI: 1.05–35.2, *P* < 0.05), HD (OR: 14.2, 95%CI: 2.10–95.8, *P* < 0.01), and smoking (OR: 17.8, 95%CI: 1.52–209, *P* < 0.05) were significantly associated with AD. The prognostic scoring model for AD within 1 year, developed through multivariate analysis, comprised the following factors: age ≥ 70 years, HD, smoking, and CONUT score ≥ 5. The probability of AD within 1 year was predicted using the following formula, where each factor was substituted with 1 if present and 0 if not: Probability of AD within 12 months = 1/{1+exp(-z)}

**Table 2.**
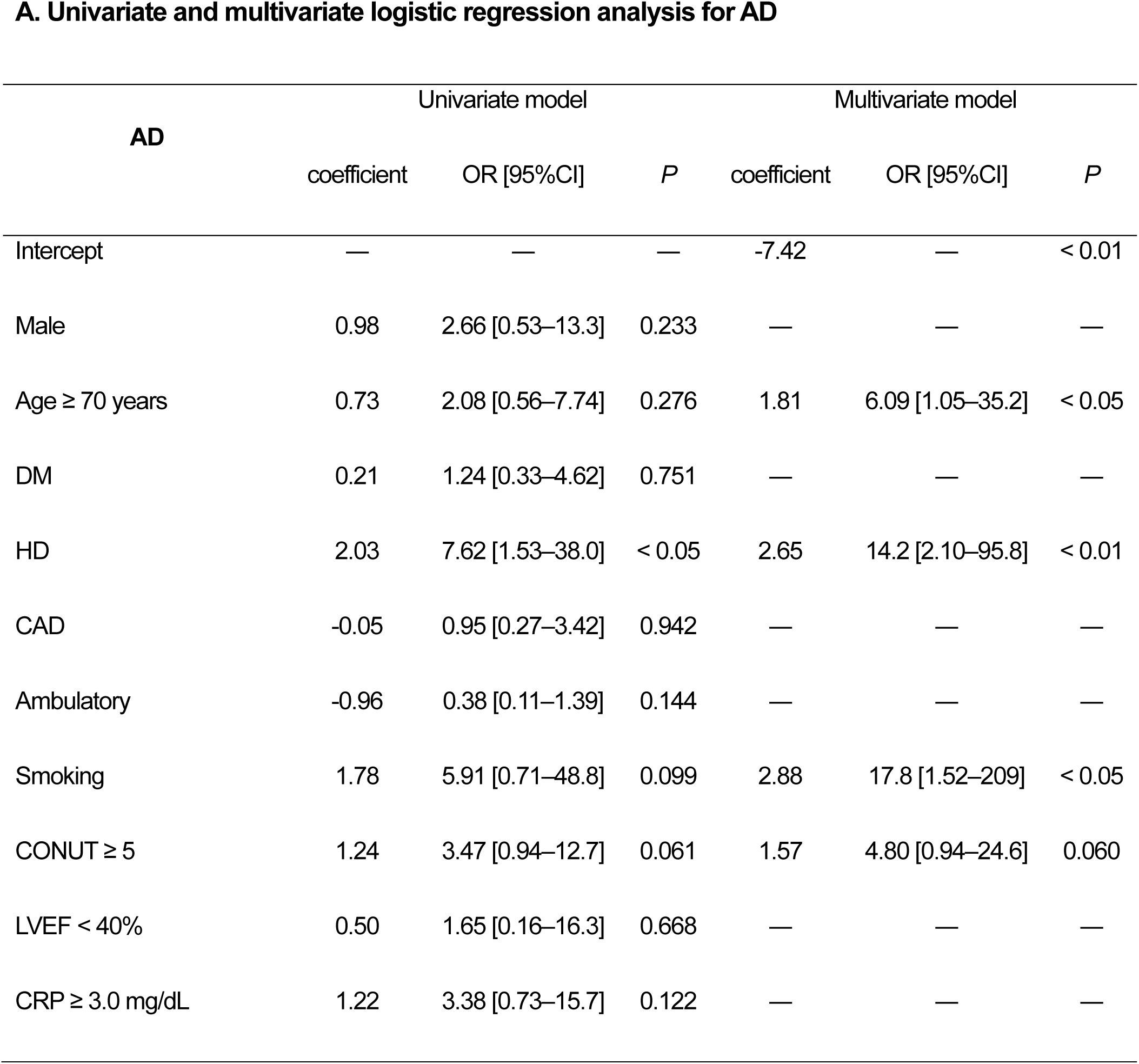
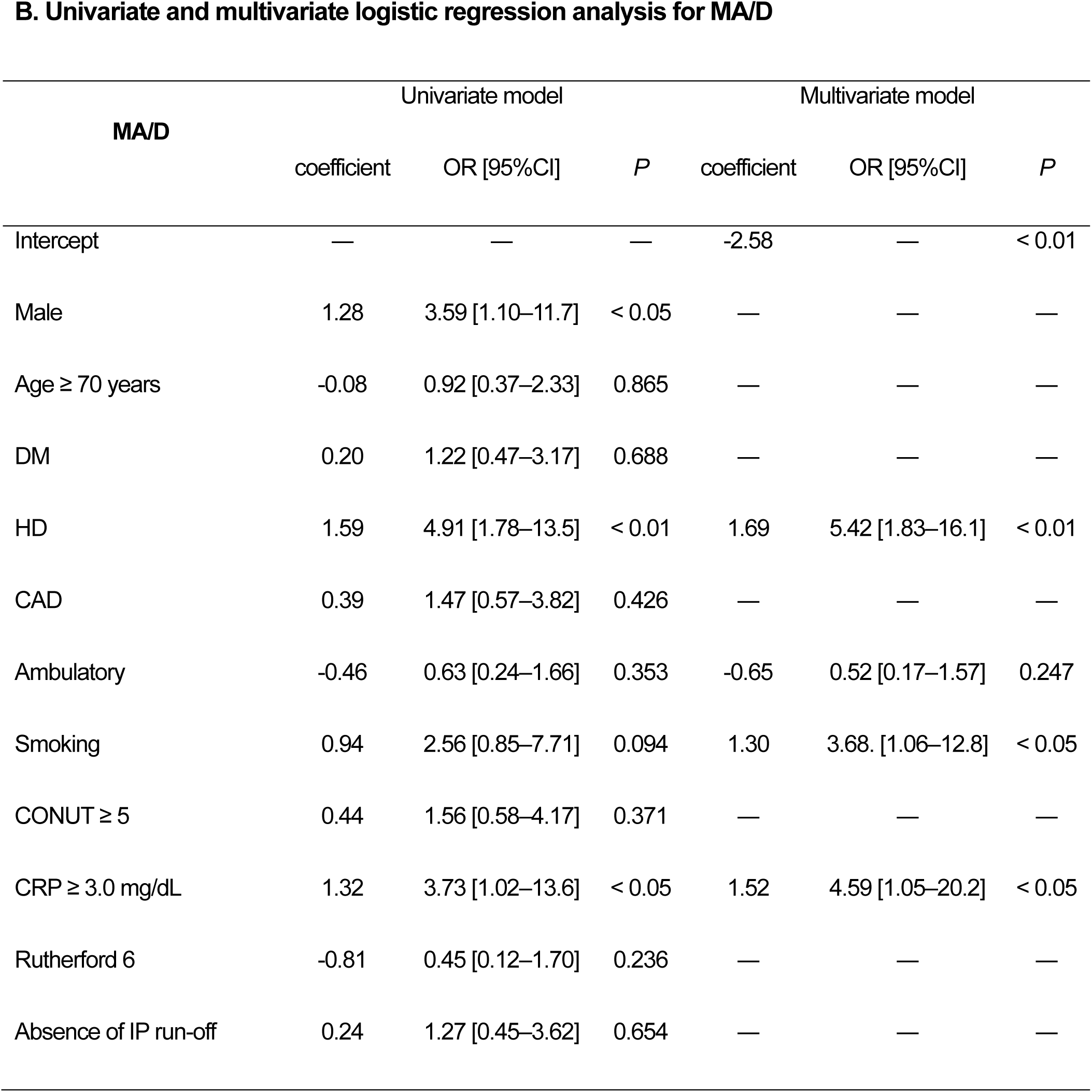
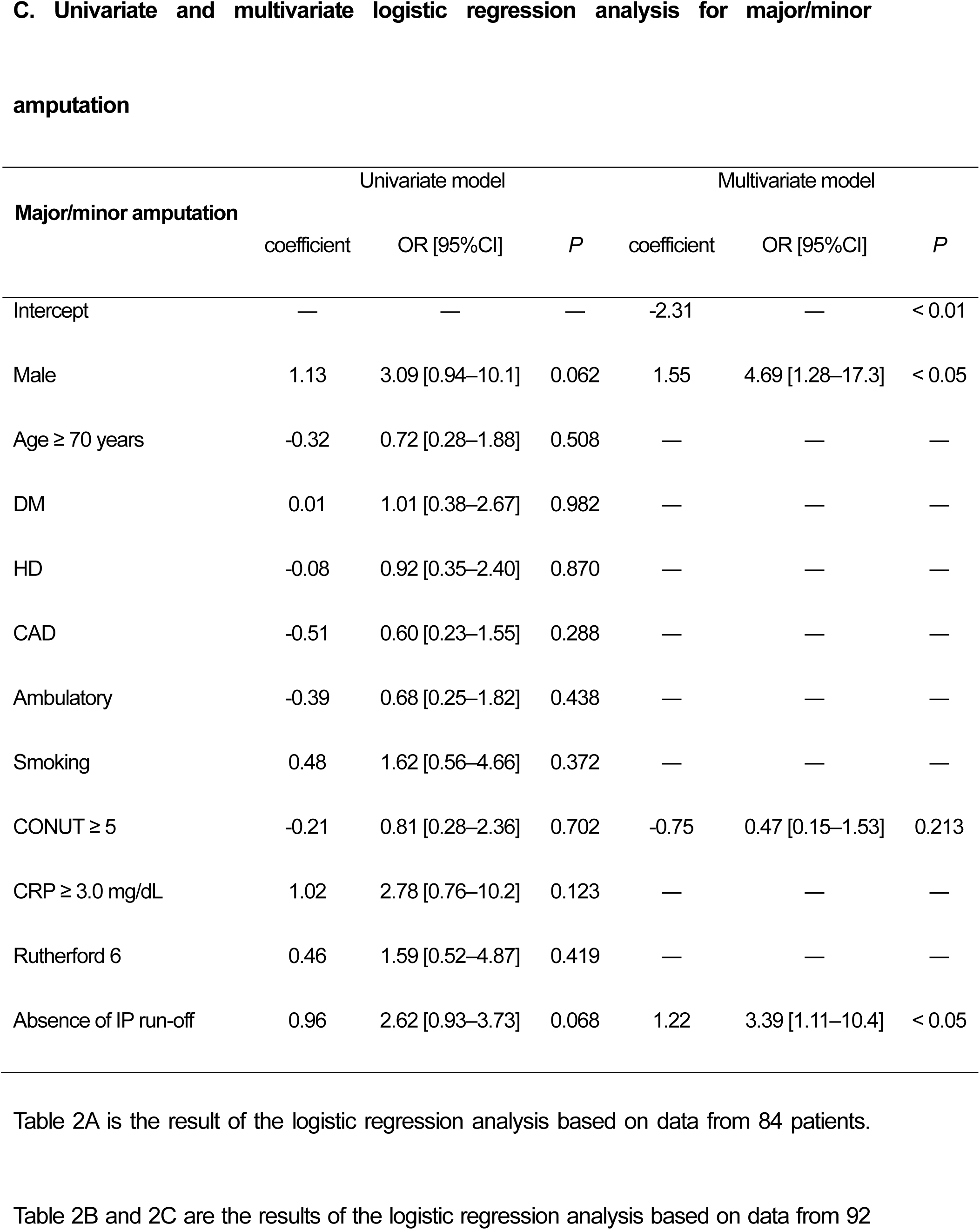
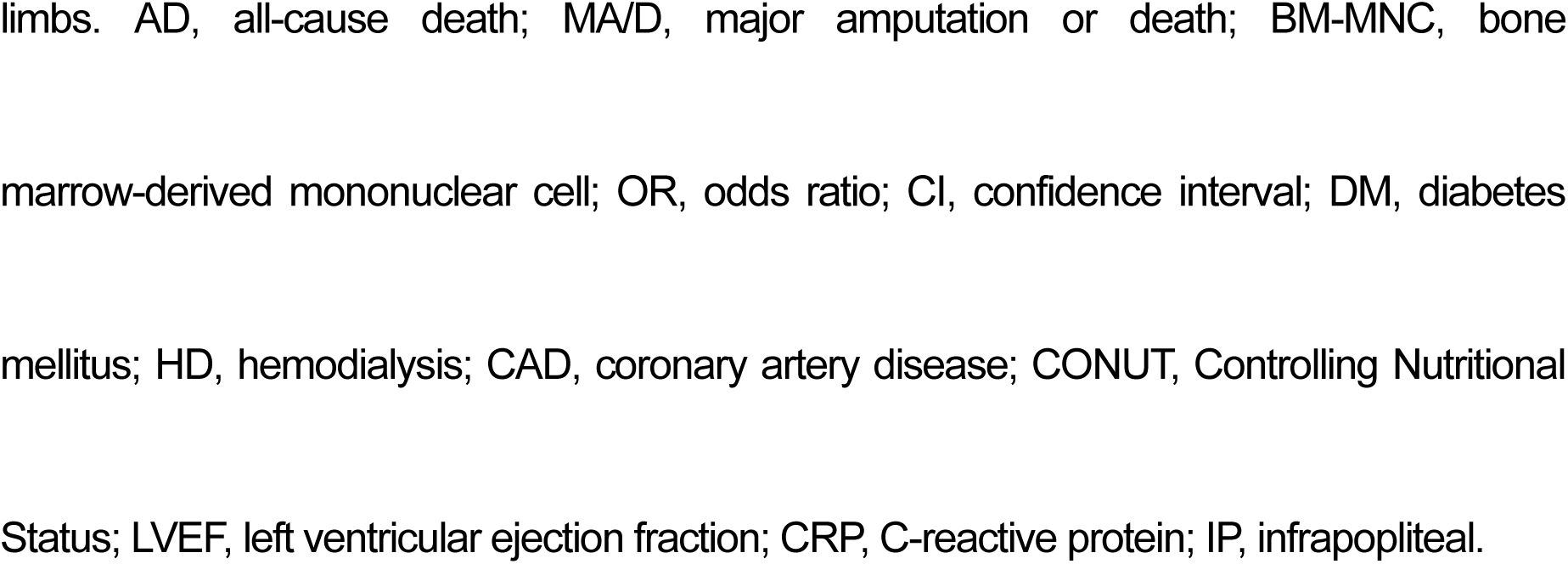
Univariate and multivariate logistic regression analysis for AD (A), MA/D (B), and major/minor amputation (C) within 1 year after BM-MNC implantation.

Where:

z = 1.81 [Age ≥ 70] + 2.65 [HD] + 2.88 [Smoking] + 1.57 [CONUT ≥ 5] – 7.42

This formula served as the basis for determining the score “y” using the following equation:

y = [Age ≥ 70] + 2 [HD] + 2 [Smoking] + [CONUT ≥ 5]

The area under the curve (AUC) of the receiver operating characteristic (ROC) curve was 0.87, and the sensitivity and specificity for AD within 1 year were 91% and 73%, respectively, when y was ≥ 4 (Supplementary Figure S1A). This scoring model was used to divide the 84 patients included in the patient data into Groups 1 (y < 4) and 2 (y ≥ 4). Figure 4A shows the long-term prognosis of both groups. The respective OS rates after 6 months, 1 year, 5 years, and 10 years were 98.2%, 96.2%, 64.9%, and 35.8% in Group 1 (n=54) and 76.7%, 69.7%, 23.2%, and 23.2% in Group 2 (n=30). A significant difference was observed in the long-term prognoses of the two groups (log-rank test, *P* < 0.01).

**Figure 4.**
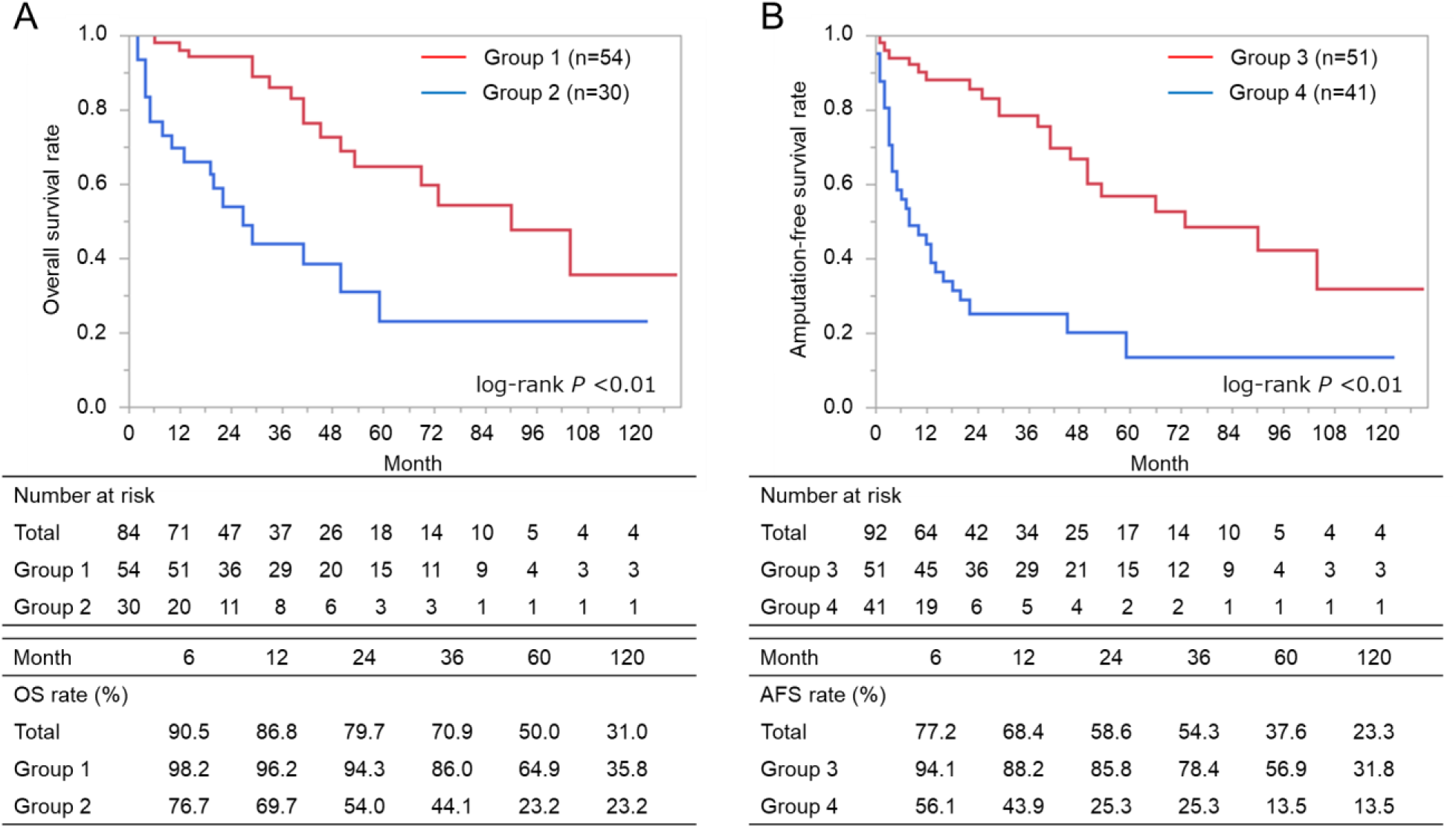
Comparison of two groups divided by scoring models. Figure 4A shows a significant difference in the OS rate between Group 1 and Group 2 (log-rank *P* <0.01). Figure 4B reveals a significant difference in the AFS rate between Group 3 and Group 4 (log-rank *P* <0.01). These groups were determined by the prognostic scoring model for the probability of AD and MA/D within 1 year, developed through each multivariate analysis. OS, overall survival; AFS, amputation-free survival; AD, all-cause death; MA/D, major amputation or death.

Variables related to MA/D within 1 year were similarly analyzed (Table 2B). Univariate analysis showed that male sex (OR: 3.59, 95% CI: 1.10–11.7, *P* < 0.05), HD (OR: 4.91, 95% CI: 1.78–13.5, *P* < 0.01), and C-reactive protein (CRP) levels ≥ 3.0 mg/dL (OR: 3.73, 95% CI: 1.02–13.6, *P* < 0.05) were significantly associated with MA/D. Stepwise multivariate analysis showed that HD (OR: 5.42, 95% CI: 1.83–16.1, *P* < 0.01), smoking (OR: 3.68, 95% CI: 1.06–12.8, *P* < 0.05), and CRP levels ≥ 3.0 mg/dL (OR: 4.59, 95% CI: 1.05–20.2, *P* < 0.05) were significantly associated with MA/D. The prognostic scoring model for MA/D within 1 year, developed through multivariate analysis, included the following factors: HD, ambulatory ability, smoking, and CRP levels ≥ 3.0 mg/dL. The probability of MA/D within 1 year could be predicted using the following formula, where each factor was substituted with 1 if present and 0 if not: Probability of MA/D within 12 months = 1/{1+exp(-z)}

Where:

z = 1.69 [HD] – 0.65 [Ambulatory] + 1.30 [Smoking] + 1.52 [CRP ≥ 3.0] – 2.58

This formula served as the basis for determining the score “y” using the following equation:

y = 5 [HD] – 2 [Ambulatory] + 4 [Smoking] + 5 [CRP ≥ 3.0]

The AUC of the ROC curve was 0.80, and the sensitivity and specificity for MA/D within 1 year were 80% and 70%, respectively, when y was ≥ 5 (Supplementary Figure S1B). This scoring model was used to divide the 92 affected limbs included in the limb data into Groups 3 (y < 5) and 4 (y ≥ 5). Figure 4B shows the long-term prognosis of both groups. The respective AFS rates after 6 months, 1 year, 5 years, and 10 years were 94.1%, 88.2%, 56.9%, and 31.8% in Group 3 (n=51) and 56.1%, 43.9%, 13.5%, and 13.5% in Group 4 (n=41). A significant difference was observed in the long-term prognoses of the two groups (log-rank test, *P* < 0.01).

Table 2C shows the results of logistic regression analysis of variables related to major/minor amputations within 1 year of BM-MNC implantation as secondary endpoints. In univariate analysis, no significantly associated variables were found, including male sex (OR: 3.09, 95% CI: 0.94–10.1, *P* = 0.06), HD (OR: 0.92, 95% CI: 0.35–2.40, *P* = 0.87), smoking (OR: 1.62, 95% CI: 0.56–4.66, *P* = 0.37), CONUT score ≥ 5 (OR: 0.81, 95% CI: 0.28–2.36, *P* = 0.70), CRP levels ≥ 3.0 mg/dL (OR: 2.78, 95% CI: 0.76–10.2, *P* = 0.12), Rutherford category 6 (OR: 1.59, 95% CI: 0.52–4.87, *P* = 0.42), and absence of infrapopliteal (IP) run-off (OR: 2.62, 95% CI: 0.93–7.37, *P* = 0.07). Stepwise multivariate analysis showed that male sex (OR: 4.69, 95% CI: 1.28–17.3, *P* < 0.05) and absence of infrapopliteal run-off (OR: 3.39, 95% CI: 1.11–10.4, *P* < 0.05) were significantly associated with major/minor amputations.

Table 3 shows the results of univariate logistic regression analysis using the counts of BM-MNCs and CD34+ cells as variables related to AD and MA/D within 1 year of BM-MNC implantation. The BM-MNC count (per 10^9^), BM-MNC count per body weight (per 10^7^/kg), CD34+ cell count (per 10^7^), and CD34+ cell count per body weight (per 10^5^) were all significant prognostic factors associated with a reduced incidence of AD. Similarly, the BM-MNC count (per 10^9^) and BM-MNC count per body weight (per 10^7^/kg) were significant prognostic factors associated with a reduced incidence of MA/D.

**Table 3.** Univariate logistic regression analysis for AD and MA/D associated with BM-MNC and CD34+ cell counts.

| **BM-MNC and CD34+**<br>**cell counts** | **AD within 1 year** |  |  | **MA/D within 1 year** |  |  |
| --- | --- | --- | --- | --- | --- | --- |
|  | coefficient | OR [95%CI] | *P* | coefficient | OR [95%CI] | *P* |
| BM-MNC, per 10<sup>9</sup> | -1.41 | 0.24 [0.08–0.71] | < 0.01 | -0.94 | 0.39 [0.20–0.76] | < 0.01 |
| BM-MNC/BW, per 10<sup>7</sup>/kg | -0.58 | 0.56 [0.33–0.94] | < 0.05 | -0.56 | 0.57 [0.40–0.82] | < 0.01 |
| CD34+ cell, per 10<sup>7</sup> | -0.68 | 0.51 [0.29–0.89] | < 0.05 | -0.14 | 0.87 [0.69–1.08] | 0.207 |
| CD34+/BW, per 10<sup>5</sup>/kg | -0.32 | 0.73 [0.54–0.97] | < 0.05 | -0.11 | 0.90 [0.79–1.02] | 0.089 |

The results of AD and MA/D were based on data from 84 patients and 94 limbs, respectively. AD, all-cause death; MA/D, major amputation or death; BM-MNC, bone marrow-derived mononuclear cell; CD34+, CD34 surface antigen positive; BW, body weight; CD34+, CD34 surface antigen positive; OR, odds ratio; CI, confidence interval.

### Safety

Table 4 presents information on adverse events that occurred within 6 months of BM-MNC implantation. AD, CAD, CVD, and heart failure were observed in 8 (10%), 4 (5%), 2 (2%), and 3 (4%) patients, respectively, with no cases of aortic disease among the patients. MA/D, major amputation, minor amputation, revascularization, and major/minor amputation were observed in 20 (21%), 12 (13%), 9 (10%), 14 (15%), and 18 (20%) limbs, respectively. No direct causal relationship existed between any of the adverse events and BM-MNC implantation.

**Table 4.** Adverse events during 6 months of follow-up.

| **Adverse events from patient data** | **N=84** |
| --- | --- |
| AD | 8 (10) |
| HF | 3 (4) |
| CAD | 4 (5) |
| CVD | 2 (2) |
| Aortic disease | 0 (0) |
|  |  |
| **Adverse events from limb data** | **N=92** |
| MA/D | 20 (21) |
| Major amputation | 12 (13) |
| Minor amputation | 9 (10) |
| Revascularization | 14 (15) |
| Major/minor amputation | 18 (20) |

“Patient data” and “limb data” are based on data from 84 patients and 92 limbs, respectively.
AD, all-cause death; HF, heart failure; CAD, coronary artery disease; CVD cerebrovascular disease; MA/D, major amputation or death.

## DISCUSSION

Patients with CLTI often face elevated rates of limb amputation and mortality. The most important goal of treatment for patients with CLTI is to prevent limb amputation and enhance their life expectancy. The first-line treatment to achieve this goal is revascularization, encompassing EVT and surgical bypass. In a recent Japanese study, the AFS rate at 3 years after revascularization was 52% for both EVT and surgical bypass (SPINACH registry) [8]. This means that many patients with CLTI remain at considerable risk of limb amputation and mortality despite receiving conventional treatments, including revascularization.

For these patients with no-option CLTI, therapeutic angiogenesis using BM-MNC implantation is a potential recourse. Multiple studies have shown the effectiveness and safety of BM-MNC implantation [13–23]. In 2008, Matoba et al. reported that the OS and AFS rates at 3 years after BM-MNC implantation were 80% and 60%, respectively, in 74 patients with no-option CLTI resulting from atherosclerotic LEAD (TACT follow-up study) [17]. A 2011 report by Idei et al. indicated that the AFS rate at 4 years in 25 patients with atherosclerotic LEAD-derived no-option CLTI who underwent BM-MNC implantation was significantly higher than that in 30 patients who did not undergo BM-MNC implantation (48% vs 0%, *P* < 0.0001) [18]. In a long-term prognosis analysis performed by Kondo et al., 374 patients with no-option CLTI due to atherosclerotic LEAD had OS rates at 1, 5, and 10 years after BM-MNC implantation of 95.6%, 87.3%, and 68.6%, respectively, and AFS rates of 82.7%, 71.8%, and 59.1%, respectively [20]. In addition, BM-MNC implantation has been reported to improve various indicators of limb ischemia, including the resting pain, pain-free walking distance, ulcer size, and transcutaneous partial pressure of oxygen (tcPO_2_) [13–17, 22, 23].

As demonstrated in previous studies, therapeutic angiogenesis using BM-MNC implantation is effective for no-option CLTI; however, the long-term prognosis and ischemic indicators of no-option CLTI resulting from atherosclerotic LEAD appear to be inferior to those resulting from TAO or CVD. This is probably because patients with atherosclerotic LEAD tend to be older, exhibit a higher prevalence of CAD, experience more severe complications, and have a poorer nutritional status than those with TAO and CVD [13, 17, 18, 20]. In patients with CLTI receiving conventional treatment, various prognostic factors have also been reported, including age, activity of daily living (ADL), HD, heart failure (or low left ventricular ejection fraction), and Rutherford classification [5, 24–26]. However, in patients with no-option CLTI after BM-MNC implantation, prognostic factors are poorly understood. According to the results of the logistic analysis conducted in the present study, an age ≥ 70 years, HD, and smoking were prognostic factors that influenced the 1-year prognosis for the OS rate in patients with atherosclerotic LEAD-induced no-option CLTI. Factors that influenced the 1-year prognosis for the AFS rate were male sex, HD, smoking, and CRP levels ≥ 3.0 mg/dL. A scoring formula, developed through multivariate analysis, was then used to effectively identify a group with a favorable long-term prognosis with respect to both the OS and AFS rates. In the present study, the OS rates at 1 year, 5 years, and 10 years after BM-MNC implantation were 86.8%, 50.0%, and 31.0%, respectively. Additionally, the AFS rates at 1, 5, and 10 years were 68.4%, 37.6%, and 23.3%, respectively. In the favorable prognostic groups obtained by the formulas described above, the OS rates at 1, 5, and 10 years were 96.2%, 64.9%, and 35.8%, respectively, while the AFS rates were 88.2%, 56.9%, and 31.8%, respectively. Although further studies are desired to evaluate the accuracy of these formulas, they may help predict which patients are likely to benefit from BM-MNC implantation before treatment.

The present study revealed that the BM-MNC and CD34+ cell counts had an important relationship with AD and MA/D 1 year after BM-MNC implantation. A randomized trial reported that the BM-MNC dose was an independent predictor of ulcer healing (PROVASA trial) [27]. BM-MNCs from patients with CAD have a decreased ability to form new blood vessels [28]. It is unclear how BM-MNCs and CD34+ cells are related to patient characteristics such as sex, age, and nutritional status. However, the cell counts may be useful for predicting the prognosis following BM-MNC implantation.

Global vascular guidelines advocate for a revascularization approach based on the Global Limb Anatomic Staging System, which reflects the complexity of anatomic vascular lesions [1]. Notably, establishing a direct flow into the pedal arch has shown significant promise in improving limb salvage rates and AFS [29, 30]. In this study, the multivariate logistic analysis showed that the absence of IP run-off and male sex were significant prognostic factors of major/minor amputation. This finding suggests that revascularization of IP arteries before BM-MNC implantation might prevent limb amputation. Therefore, a combined approach involving therapeutic angiogenesis through BM-MNC implantation and conventional revascularization via EVT and surgical bypass holds significant potential for enhancing the prognosis of patients with no-option CLTI and improving the outlook for their affected limbs.

In summary, this study demonstrated the long-term clinical outcomes of therapeutic angiogenesis using BM-MNC implantation among Japanese patients with no-option CLTI attributed to atherosclerotic LEAD. The study also delved into the prognostic factors influencing these outcomes. The results underscore the feasibility and safety of BM-MNC implantation as a treatment modality for this patient cohort. The scoring system described in this report helps us predict the long-term prognosis of patients and their affected limbs treated by BM-MNC implantation. These results also provide valuable information for explaining the effectiveness of BM-MNC implantation to patients and their families. A prospective clinical trial for patients with atherosclerotic LEAD-induced no-option CLTI is needed to assess the effectiveness and safety of BM-MNC implantation and to evaluate the impact of each factor on the prognosis.

### Study limitations

The present study had some limitations. This was a single-center, retrospective, observational study; therefore, the possibility of selection bias and uncorrected confounding factors must be taken into consideration. Since all the patients in this study were Japanese, whether the results of this study apply to other countries or racial groups remains uncertain. Some data, such as the pain scale, pain-free walking distance, and tcPO_2_, were not adequately collected. Additionally, therapeutic angiogenesis using BM-MNC implantation for atherosclerotic LEAD-derived CLTI is not covered by national insurance in Japan, and facilities that perform the treatment are limited by law. Therefore, patients who could not afford the treatment or those in remote areas could not be enrolled. Further, this treatment is still not widely known among medical professionals in Japan, and the number of medical institutions that can provide referrals is limited. On the other hand, referrals were received from several medical institutions, and the follow-up was conducted on patients after treatment at various medical institutions. This could have led to variations in the level of treatment before and after BM-MNC implantation. Furthermore, some patients who underwent BM-MNC implantation were already scheduled for minor amputations after the treatment, which might complicate the interpretation of some results.

## Conclusion

In conclusion, this study identified prognostic factors for atherosclerotic LEAD-derived no-option CLTI, including age, hemodialysis, smoking, malnutrition, ADL, and inflammatory response, for patients or affected limbs. The scoring model that includes these factors was useful for predicting the long-term prognosis. Further research in larger and more diverse patient populations may help refine these prognostic models and improve our understanding of therapeutic angiogenesis in CLTI.

## Non-standard Abbreviations and Acronyms

AD: all-cause death
ADL: activity of daily living
AFS: amputation-free survival
AUC: area under the curve
BM-MNCs: bone marrow-derived mononuclear cells
CAD: coronary artery disease
CD34+: CD34 surface antigen-positive
CDV: collagen disease-associated vasculitis
CI: confidence interval
CLTI: chronic limb-threatening ischemia
CONUT: Controlling Nutritional Status
CRP: C-reactive protein
CVD: cerebrovascular disease
DM: diabetes mellitus
EVT: endovascular therapy
HD: hemodialysis
IP: infrapopliteal
LEAD: lower extremity artery disease
MA/D: major amputation or death
OR: odds ratio
OS: overall survival
ROC: receiver operating characteristic
TAO: thromboangiitis obliterans
tcPO_2_: transcutaneous partial pressure of oxygen
UMIN: University Hospital Medical Information Network

## Acknowledgements

The authors would like to express their gratitude for the invaluable assistance and support provided by all committee members involved in this project and members from other institutions.

## Data availability statement

All datasets generated during and/or analyzed during the current study are available from the corresponding author on reasonable request.

## Supplemental Material

**Supplementary Figure S1.**
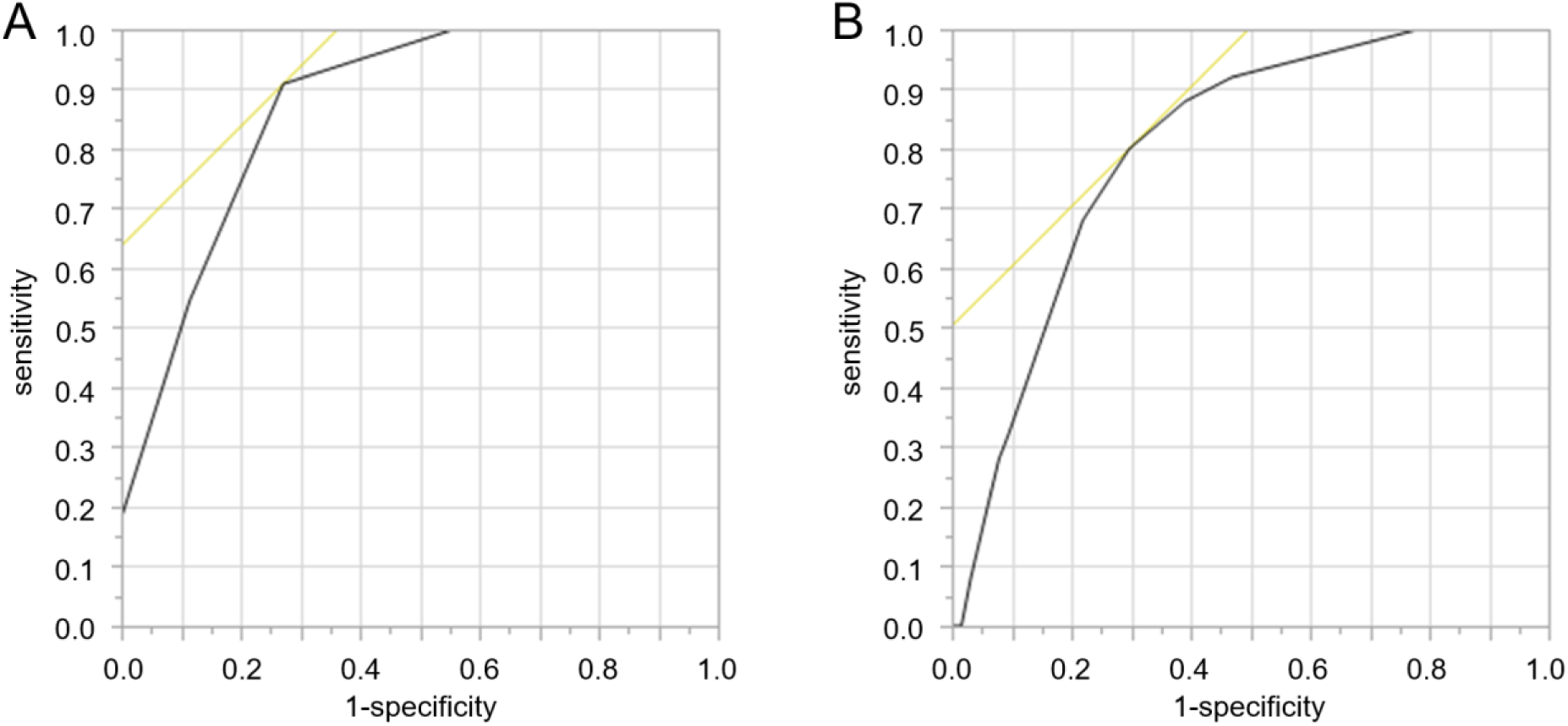
The ROC curve of the prediction model for AD (A) and MA/D. Figure S1A shows the receiver operating characteristic (ROC) curve of prognostic scoring model (included age ≥ 70 years, HD, smoking, and CONUT score ≥ 5) for AD within 1 year through multivariate analysis. The area under the curve (AUC) was 0.87. Figure S1B shows the ROC curve of prognostic scoring model (included HD, ambulatory ability, smoking, and CRP levels ≥ 3.0 mg/dL) for MA/D within 1 year. The AUC was 0.80. AD, all-cause death; HD, hemodialysis; CONUT, Controlling Nutritional Status; CRP, C-reactive protein; MA/D, major amputation or death.

